# Integrating a Non-Communicable Disease Care Cascade within Ghana’s Community-Based Health Planning and Services (CHPS) Program: the COMBINE Pilot Implementation Trial

**DOI:** 10.64898/2026.06.03.26354834

**Authors:** David J Heller, Yehia Elkersh, Engelbert A Nonterah, Irene Kuwolamo, Carol R Horowitz, Evan E Alvarez, Timothy Awine, Usha Govindarajulu, Allison P Squires, Benedict Weobong, Raymond A Aborigo

## Abstract

**Introduction:** Hypertension is the world’s leading cause of death, and depression its leading cause of disability. Control rates for these non-communicable diseases (NCDs) are low in low- and middle-income countries (LMICs). Many LMICs have programs to screen and treat underserved communities for infectious diseases, but evidence to adapt them to treat NCDs is limited. We developed and tested a non-communicable disease program through Ghana’s Community-Based Health Planning and Services (CHPS) primary care initiative.

**Methods:** We trained 8 CHPS nurses to diagnose and treat hypertension and depression through door-to-door screening and pharmacotherapy. Physician assistants provided telehealth supervision. We combined this treatment with volunteer counseling to boost medication adherence, improve mood, and change health behaviors. We called the 90-day intervention the CHPS Opportunity for Mentally and Behaviorally Integrated NCD Engagement (COMBINE).

**Results:** We recruited 60 adults from 580 screened: 37 with hypertension (mean blood pressure (BP) of 149/91 mm Hg) and 23 with depression (mean physician health questionnaire (PHQ-9) score of 13.3). After 90 days, 57/60 (95%) completed the intervention: 32/37 (86%) achieved blood pressure control (mean BP 122/75 mm Hg), and 19 of 20 (95%) achieved depression control (mean PHQ-9 score 2.0). After 12 months, 51/60 were retained: 33/37 with hypertension (89%) and 18/23 with depression (78%), with a mean BP of 121/75 and PHQ-9 score of 1.4 respectively. All 51 (100%) achieved disease control at 12 months. 5 persons left by migration and 4 by escalation to higher-level care.

**Conclusions:** The COMBINE model achieved high levels of diagnosis, care retention, and disease control, with minimal adverse events, in a remote setting with limited usual NCD care. This model suggests a novel means to improve the care cascade for these and other noncommunicable diseases through existing non-physician care models in LMICs, warranting further controlled testing at scale.

**What Is Already Known on This Topic?:** Hypertension and depression are the leading causes of global death and disability respectively. In Ghana as in many low- and middle-income countries, evidence shows rural non-physician health workers can proactively screen persons with these and other noncommunicable diseases and refer them to higher-level clinics. However, such clinics are often few and distant from rural areas, limiting access and disease control.

**What Does This Study Add?:** Our program is the first to test the hypothesis that nurses and volunteers in Ghana’s primary care program can treat hypertension and depression in remote villages - through pharmacotherapy and behavior counseling. We found that 95% of trial participants completed the 90-day intervention, and 91% achieved disease control. At 1 year, 85% remained in care, of which 100% achieved disease control. These retention and control rates meet or exceed that of higher-level health centre care.

**How Might This Study Affect Policy?:** If effective at scale in a randomized trial, this intervention could allow CHPS to markedly increase awareness, treatment, and control rates for hypertension and depression in Ghana. Its focus on promoting behaviors relevant to many non-communicable diseases - such as physical exercise and alcohol and tobacco cessation - may make it applicable to the concurrent management of multiple chronic conditions. Further, this care model could be leveraged in other health systems with a national non-physician rural health program, improving noncommunicable disease control worldwide.

## Introduction

Non-communicable diseases (NCDs) are the leading cause of human mortality. 73% of NCD deaths occur in low- and middle-income countries (LMICs).^1^ The leading cause of NCD and all-cause death globally is cardiovascular disease (CVD),^1,2^ and its leading contributor is hypertension.^3^ However, NCDs also cause most global disability, and depression is the single largest contributor.^4,5^ The burden of NCD-related death and disability is increasing rapidly in LMICs, especially in younger populations, and straining health systems that are financially and structurally unprepared to respond.^6^

Ghana is no exception: hypertension affects 34% of adults, and 43% of its deaths are due to NCDs.^7^ Both proportions are rising due to changes in lifestyle and behavior.^8,9^ Cardiovascular disease now causes 19% of all deaths.^7,10^ Only 22% of persons with hypertension receive treatment, and only 6% achieve blood pressure control.^11^ The adult prevalence of depression is 16% to 25%,^12,13^ but less than 2% of cases are detected,^13^ and even fewer achieve disease control - with no differences across gender.^11-13^

Drivers of hypertension in Ghana include alcohol use, physical inactivity, and obesity.^14^ These behaviors are exacerbated by depression in turn.^9,15^ Detection and control of these NCDs in Ghana include a limited health workforce to screen and treat them: about 2.7 physicians per 10,000 persons in 2023,^16^ relative to an ideal of 10.^17^ There is an overall staff shortfall of 41% within the Ghana Health Service, with wider gaps in rural areas.^18^

Fortunately, the Ghana Health Service has a robust primary care system designed to close health access gaps in rural preventive and curative primary care. The CHPS initiative, created and refined in the 1990s in Navrongo in Ghana’s Upper East Region, trained nurses to improve childhood vaccination rates, household hygiene, and access to safe antenatal care, and deployed them to live and work within rural areas.^19^ CHPS nurses, called community health nurses (CHNs) proactively conduct door-to-door home visits to all households in their zones every three months to identify those needing preventive or curative care and offer it - either on the spot or at a CHPS compound. A CHPS compound is a facility within a delineated area called a CHPS zone, where primary care services are provided. Community health volunteers (CHVs) provide home education to households to reinforce nurses’ counseling messages.^19,20^

Over the past two decades, CHPS has been scaled up across Ghana, and now serves as the first tier of a primary care system backed by a health insurance scheme that has lowered child and maternal mortality.^21-23^ However, CHPS nurses and CHVs are not authorized to treat non-communicable diseases such as hypertension or depression, due to lack of evidence of efficacy or safety. Instead, CHPS nurses only screen for diseases such as hypertension, and link suspected cases to higher-level facilities (health centres or hospitals) for diagnosis and treatment by physicians and physician assistants.^24-26^

Based on the proven efficacy of CHPS,^23^ growing global evidence that nurses and other non-physicians can effectively diagnose and treat noncommunicable diseases with adequate training and supervision,^27-29^ and the increasing burden of these conditions in rural Ghana,^11,12,30^ we developed and pilot tested a program through which CHPS nurses proactively test for and treat NCDs in their zones, in lieu of health centre referral. The rationale and structure of the program emerged from formative work that showed that cardiovascular disease is a massive and growing source of death and disability in the area.^31^ We found that hypertension in particular is a source of dread, because health centers are too far away: persons referred by CHPS to make this journey act “like you cursed them or something. They don’t want to go.” ^32^ and instead “silently go back to the house and wait for death.”^33^

Subsequent qualitative research revealed widespread enthusiasm for hypertension care *within* CHPS, focused not only on medication but on patient behavior counseling around risk factors such as alcohol and tobacco use, and depressed mood.^32-34^ CHPS nurses felt comfortable treating uncomplicated hypertension if adequately trained, but advised volunteers should provide the counseling because “they even know these [community] people more than us.” Depression emerged as a crucial focus of this effort because community members perceived low mood as a direct cause of hypertension and CVD,^34,35^ as well as a cause and an effect of behaviors like alcohol use and sedentary behavior that further exacerbate the growing CVD burden.^34,35^ Persons of all genders echoed this view in our formative research.^31-33^ We therefore designed and pilot-tested an integrated hypertension and depression intervention through CHPS called COMBINE (CHPS Opportunity for Mentally and Behaviorally Integrated NCD Engagement.^36^ Our hypothesis was that CHPS nurses and CHVs could effectively screen for, diagnose, and treat uncomplicated hypertension and depression at the community level with telehealth support.

## Methods

### Study Design, Setting, and Approach

We conducted an uncontrolled pilot trial of the effectiveness, implementation feasibility, and safety of COMBINE in two districts in the Upper East Region of Ghana, called Kassena-Nankana East and West (Figure 1). The districts are rural, low-income and populated chiefly by subsistence farmers.^37^ Some 70% of the community has no formal education; 11% are smokers; and 20% use alcohol problematically.^38^ Approximately 25% of adults in the Upper East live with hypertension and over 30% with depression,^30,39^ with no significant difference between genders – despite greater smoking prevalence in men and alcohol misuse in women.^30,31,38^ The community is roughly equally divided between members of the Kassena and Nankani ethnic groups.^37^ CHPS coverage is almost universal across all non-urban areas of the two districts: outside the main cities of Navrongo and Paga, every community and residence lies within a CHPS zone, staffed with two or more salaried CHNs aided by four or more unpaid CHVs appointed by the community.^24^

**Figure 1:**
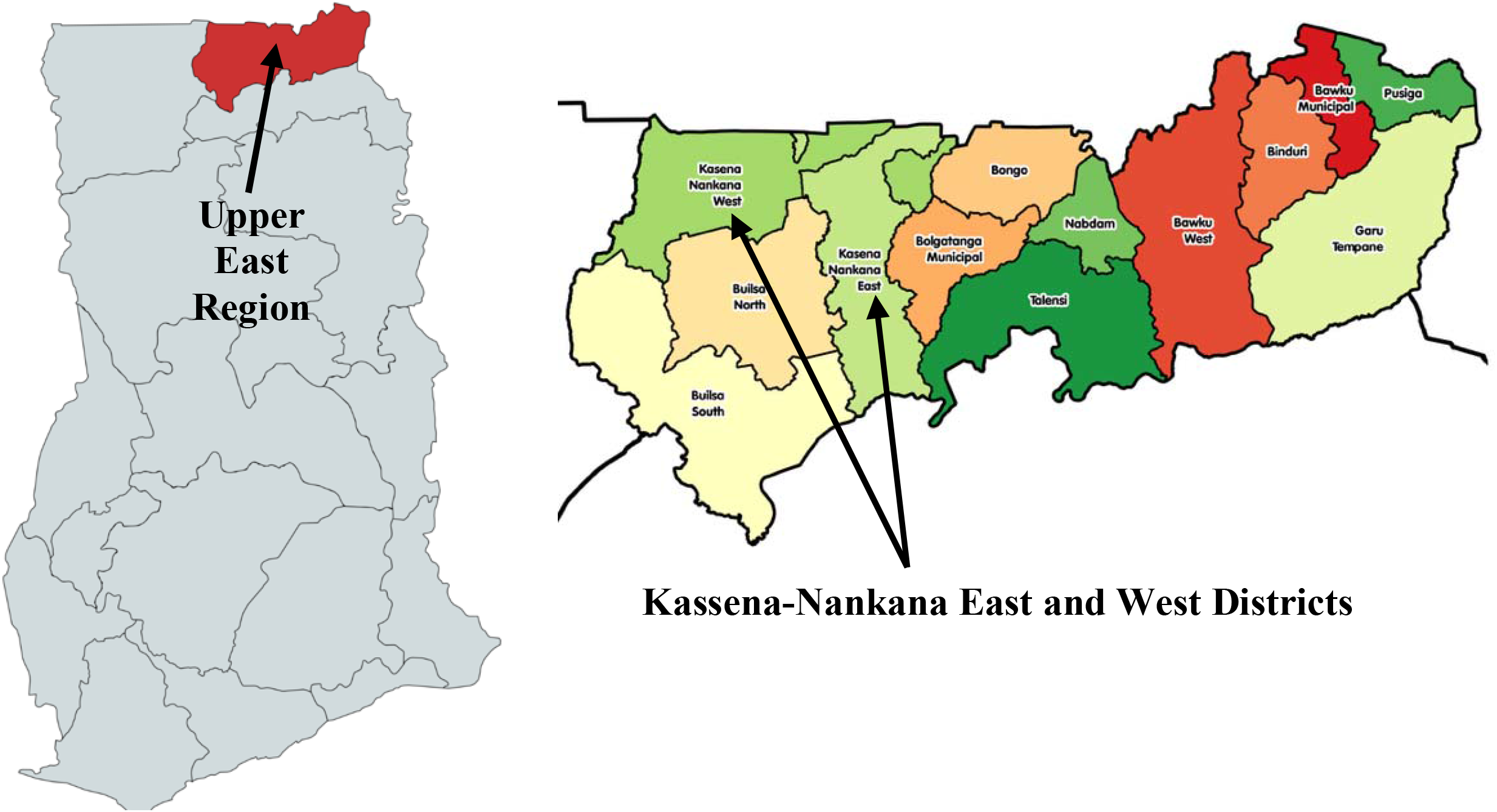
Map of the Kassena-Nankana Districts of Ghana.

We selected four CHPS zones in which to implement the intervention: Yua, Navio, Wuru, and Mirigu. The selection aimed to achieve an even geographic distribution across the districts and a balance of persons from Kassena and Nankani communities.^37^ We recruited two CHPS nurses in each of the four zones, and used their input to select two CHVs in each zone. We sought CHVs whom nurses felt were hardworking; who were literate in oral and written English in addition to local languages; and who were amenable to providing behavior counseling to persons with hypertension and/or depression.

### Recruitment and Eligibility Criteria

We directed each nurse to screen all community members aged 35-70 in their zone for hypertension and depression as an additional component of their usual quarterly door-to-door wellness visits to each household in the zone.^24^ We selected this age range because atherosclerotic heart diseases are uncommon in persons under 35,^40^ and because guidelines for management of these conditions change in persons over 70 years due to reduced life expectancy.^41,42^ We included persons of both male and female sex and all gender identities, because these NCDs are equally prevalent across genders in Ghana.^11-13^

Based on prior qualitative evaluation of CHPS nurses’ clinical skill level,^32,33^ we enrolled only persons with *uncomplicated* hypertension, (BP of 140-179 mm Hg systolic or 90-99 mm Hg diastolic pressure), due to the high risk of acute atherosclerotic events or death with higher pressures (e.g., hypertensive urgency). For depression, we targeted *moderate* disease as measured by a Physician Health Questionnaire (PHQ) 9-question survey: over 10 but under 20.^43,44^ All participants received a PHQ-2 screening survey, and those scoring over a minimum threshold of 1 received a PHQ-9. The PHQ-9 is used and validated in global settings, including northern Ghana.^43,44^ We excluded all persons who self-reported prior atherosclerotic disease such as heart attack or stroke, due to elevated risk of adverse events and lower BP goals. We also excluded persons who reported symptoms of psychosis, anxiety, bipolar disorder, epilepsy, or suicidality per a standardized questionnaire. We referred persons with any of these conditions to higher-level health centres given the complexity of their management.^45-47^ Lastly, we excluded persons with diabetes, pregnancy, or other unexpected findings on urinalysis below.

### Intervention Protocol

For blood pressure, persons were first screened in a sitting position with the cuff at heart level on the left arm. All cuffs were calibrated and sized appropriately for age. Persons with an elevated initial blood pressure received two blood pressure rechecks, and the average of these results was deemed the final result. For depression, a single screening PHQ-2 and subsequent PHQ-9 test result was sufficient. Persons meeting the criteria above on home screening were invited to come to the CHPS compound within two days. There, persons with elevated blood pressure received a further recheck to confirm the diagnosis. Persons who met inclusion criteria for either condition underwent an oral survey for the exclusion criteria above. They then underwent a random blood glucose check and a urinalysis. We excluded persons with a random blood glucose of 200 mg/dL (11.1 mmol/L), or any evidence of proteinuria on urinalysis. Persons of female sex received a urine pregnancy test and were excluded if positive. We offered enrollment to all persons screening negative for the above. All persons consenting to enroll were separately examined by a study physician to confirm clinical stability and exclude unrecognized comorbidities.

Enrolled participants received seven weekly home visits by a community health volunteer over two to three months, providing counseling on five elements of behavior change common to multiple NCDs: medication and treatment adherence; tobacco and alcohol reduction; a diet rich in fruits and vegetables and low in salt and fat; regular physical exercise; and activities and behaviors to improve mood. These were based on principles of motivational interviewing (MI) and behavioral activation (BA),^48.49^ both of which have proven impact on behavior change.^50,51^ They can be delivered by laypersons if appropriately trained,^52,53^ and can be integrated to address both depression and chronic disease self-management.^54,55^ Participants were separately invited to visit their CHPS nurse biweekly for a total of four medication and disease tracking (blood pressure and/or PHQ-9 score) visits.

Persons with hypertension were offered amlodipine 5 mg at study enrollment, due to its efficacy in hypertension control and safety for use without laboratory monitoring. Thereafter, the CHPS nurse monitored blood pressure at four biweekly meetings over the following two to three months, and if blood pressure was uncontrolled, increased medication in a stepwise fashion according to a locally-modified version of the HEARTS protocol,^56^ starting with amlodipine 10 mg, then lisinopril 20 mg, then lisinopril 40 mg, then bendroflumethiazide 20 mg. Persons with depression were to receive medication only if their PHQ-9 score did not decrease below 10 at the third visit with counseling alone: either amitriptyline 25 mg (due to ready availability in the health system) or fluoxetine 10 mg (in persons over 50 or with evidence of cardiovascular disease) per the WHO MHGap protocol.^57^ CHPS nurses obtained express permission, via a secure electronic correspondence system, from a physician assistant at their local health centre, to initiate or titrate any medication above. We used this protocol because only health centre physician assistants and physicians can dispense or titrate these medications per current Ghana Health Service protocols.

Following this intervention, we measured systolic blood pressure and PHQ-9 score at 90 days as the primary trial endpoint. We selected this time period due to evidence that rapid blood pressure and/or depression control achieves superior outcomes.^58^ However, in order to evaluate maintenance of intervention effect, we continued to follow participants every 90 days thereafter, offering quarterly CHV and nurse visits and measuring systolic blood pressure, PHQ-9 score, and other below endpoints at these intervals. See **Figure 1** for a summary of the care protocol.

### Training and Implementation

Study nurses and volunteers were trained in-person in March of 2022. Nurse training focused on screening (e.g., accurate blood pressure measurement), enrollment, and treatment protocols as well as escalation scenarios for ill patients. Health centre physician assistants attended these training sessions as well, to understand and supervise the care protocols and aid CHPS nurses in managing adverse events. CHV trainings taught the precepts of motivational interviewing and behavioral activation; how to adapt the focus of counseling to specific participant needs (e.g., alcohol misuse); and how to elicit participant behavior change (per MI) and set goals to achieve it (per BA).^48,49^ We trained CHVs to 1) help patients identify what behaviors to change, 2) set an action plan for change, and 3) identify specific goals by week for change, through the SMART framework for feasibility.^59^ Trainees were evaluated through role-play scenarios and feedback. Trial recruitment and launch began in April 2022 and the intervention ended in August 2023.

**Figure 2:**
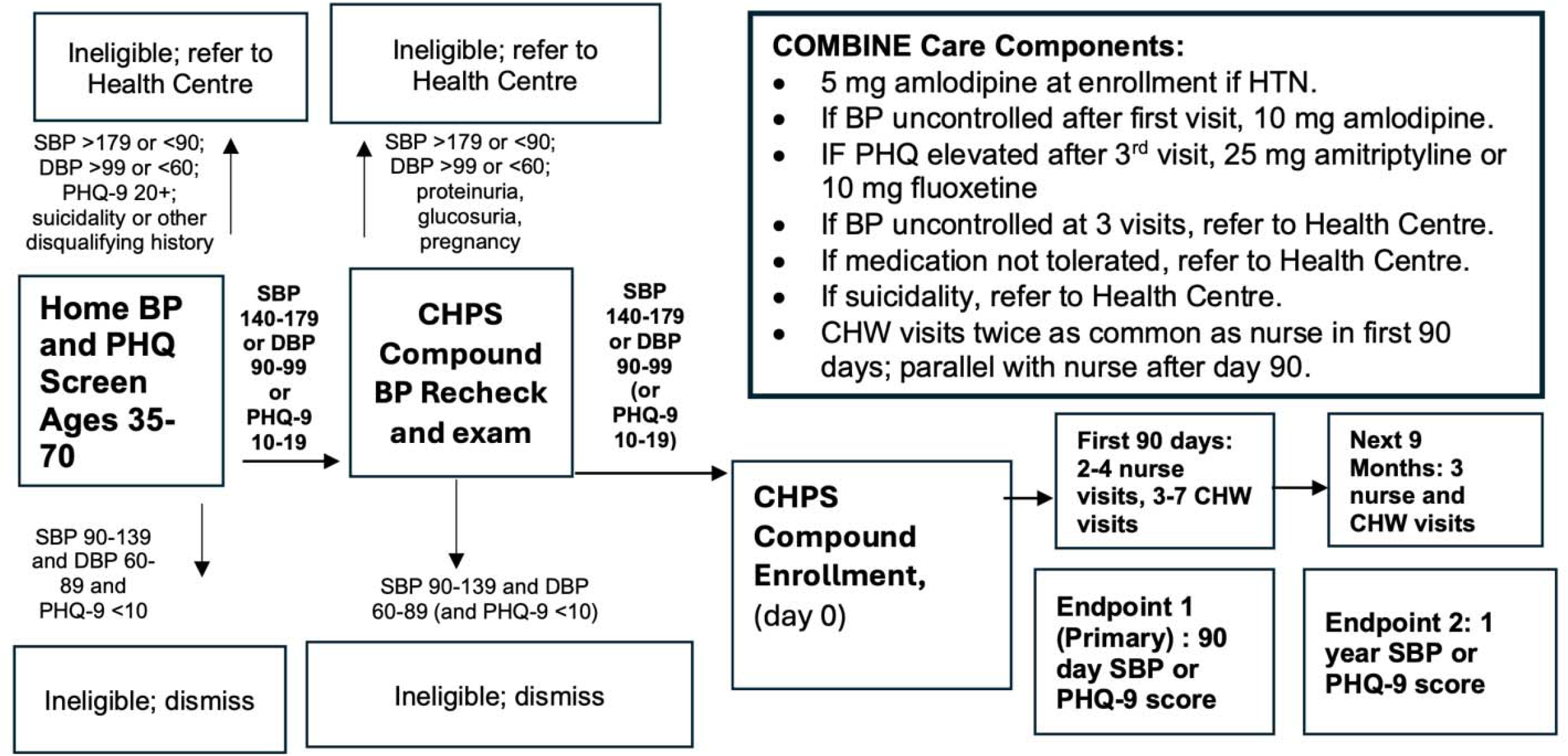
COMBINE Care Protocol.

### Data Capture and Analysis

CHPS nurses captured blood pressure and PHQ-9 scores, weekly alcohol and tobacco use, physical exercise minutes (daily and weekly), and age and gender data on all persons screened for the intervention, as well as demographic data such as marital status, number of children, occupation, religion, and ethnicity. They also captured data on who met exclusion criteria above such as prior heart attack or pregnancy, plus height, weight, and vital signs as documented by the study physician. At each CHV home visit, we documented self-reported medication adherence, tobacco and alcohol use, and SMART goals and plans to improve these outcomes (e.g., reduction in weekly alcoholic drinks consumed). Then, at each CHPS compound visit, we documented PHQ-9 and blood pressure, other vital signs, medications given, medication side effects noted, reinforcement discussions on SMART goals, and any adverse events requiring discussion or triage with the physician assistant or study physician. At the two primary study endpoint dates of 90 days and 1 year, we again checked blood pressure and PHQ-9 as well as alcohol, tobacco, and physical exercise. For those on medication, we further queried self-reported medication adherence using the widely-validated Adherence to Medications and Refills (ARMS) scale.^60^ We analyzed these data with respect to descriptive outcomes (mean, standard deviation, minimum and maximum), and compared differences by chi-squared test (for categorical variables) and a two-sided two-sample t-test, Wilcoxon rank-sum test, or Wilcoxon signed-rank test (for continuous variables, normally or non-normally distributed). We set a significance level of 0.05 in all cases. We performed all analyses in Stata.^61^.

### Patient and Public Involvement

Patients and community members have engaged with this research since its inception. Formative work informing intervention design expressly elicited these perspectives,^32-35^ and this input drove our decision to focus on mental as well as cardiovascular health as an intervention and outcome,^34,35^ in addition to program design and implementation decisions such as the use of one-on-one rather than group counseling.^33-35^ Community leader discussions were used to assess acceptability of the program and community willingness to participate. This work guided the selection of the four CHPS zones for the intervention, prioritising areas with clear expressed interest. CHVs recommended to work with were cross-checked with the community leaders before being recruited. All patients and providers recruited for the intervention were informed of the anticipated time commitment as part of the informed consent process.

### Ethical Considerations

The protocol was approved in November 2021 by the Navrongo Health Research Centre Institutional Review Board (NHRC IRB433) and the Program for the Protection of Human Subjects at the Icahn School of Medicine at Mount Sinai (PPHS 21-01138), in alignment with the principles of the Declaration of Helsinki.^62^ Participation was voluntary. Participants were told of their right to withdraw from the study at any time without suffering any consequences.

## Results

### Hypertension Screening and Recruitment

We approached 580 persons across the four study sites for home screening. Summary results appear in Figure 3 for hypertension and depression respectively. 576 of these 580 persons (99%) had a home hypertension result; 1 refused screening and 3 had an unrecorded result. 98 of 579 persons (16.9%) met study blood pressure criteria on their first blood pressure check, and 25 (4.3%) met hypertensive urgency criteria. 438 (75.6%) had a normal blood pressure and 18 (3.1%) were abnormally low. Among persons with a recorded final result, 79 were labeled as meeting study criteria; 487 had normal blood pressure; 8 had hypertensive urgency, and 2 had hypotension. However, of the 79 labeled as meeting study criteria, only 54 had three blood pressures in the study range (68%): 18 had only one elevated blood pressure but no recheck (23%); 6 (8%) had hypertensive urgency; and 1 had normal blood pressure. Similarly, among the 487 persons labeled normal, 4 (1%) fact had confirmed hypertension after 3 rechecks, 18 (4%) had an elevated initial blood pressure but no recheck, and 462 (95%) had confirmed normal blood pressure.

**Figure 3:**
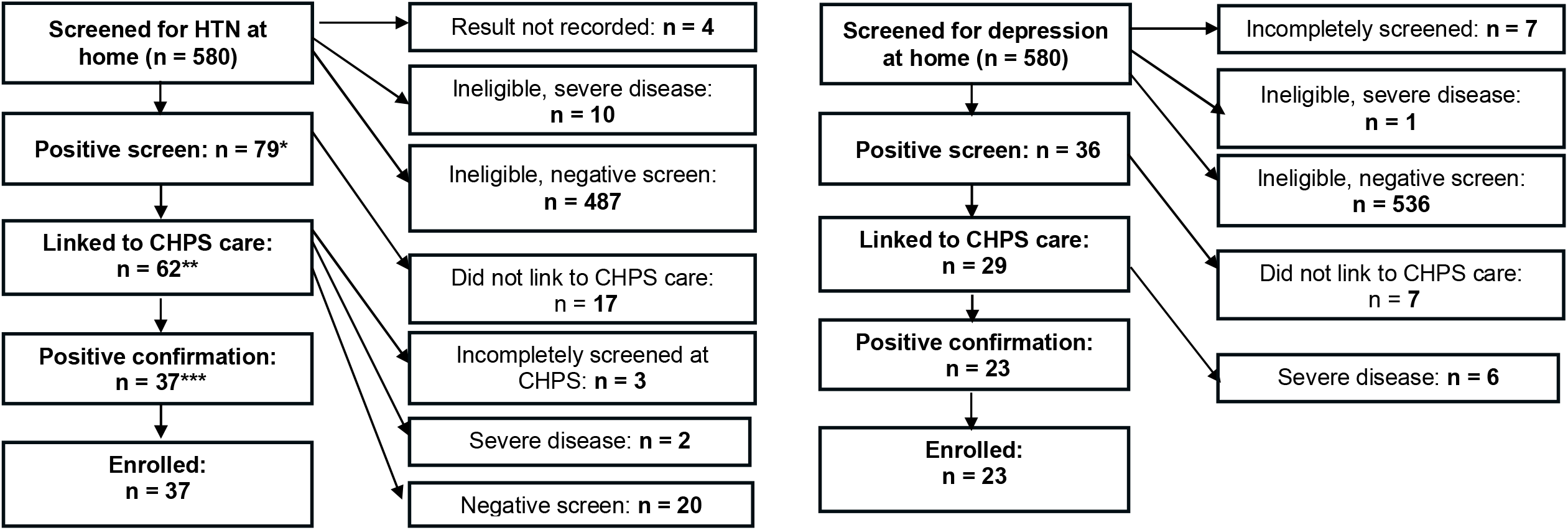
Hypertension and Depression Enrollment Cascades. *Included 54 persons with confirmed eligibility plus 25 others. **Included 45 persons with confirmed eligibility plus 17 others. ***Included 33 persons with confirmed eligibility plus 4 others.

62 persons linked to CHPS care for confirmation of hypertension status and enrollment. These included 62 of 79 (78.4%) persons labeled hypertensive and 45 of the 58 persons (77.6%) with confirmed hypertension. 37 of them (60%) were deemed eligible for the trial, and all enrolled. 33 of these persons (89%) had confirmed hypertension on CHPS screening, whereas 2 were incompletely screened and 2 had hypertensive urgency. Among the remaining 25 not enrolled, 20 (80%) had confirmed normal blood pressure at CHPS, three (12%) were incompletely screened, and 1 had hypotension and 1 had hypertensive urgency.

### Depression Screening and Recruitment

573 (98.8%) of the 580 persons were fully screened for depression. 6 refused PHQ-2 screening, whereas 1 had a PHQ-2 score above the study threshold of 3 but did not receive a PHQ-9 score. Among the 574 who received a PHQ-2 score, 415 had a score of zero; 121 had a score of 1 or 2; and 38 had a score of 3 or more. To boost enrollment, the trial lowered the threshold for follow-up PHQ-9 screening from 3 to 1 after enrollment began - as a result, 37 of 38 persons with a PHQ-2 of 3 or more and 92 of 121 persons with a PHQ score of 1 or 2 received a PHQ-9, for a total of 129 people. Among those 129, none had a PHQ-9 score of zero; 45 had a score of 1-<5 (no depression), 47 scored between 5 and<10 (mild depression), 36 scored 10-<20 (moderate depression, eligible for trial) and 1 scored 20 or more (severe depression).

29 of these persons linked to CHPS follow-up depression screening, all of whom met moderate depression criteria (80.6% of those eligible). These persons each underwent further evaluation for evidence of comorbid mental illness, of which six met criteria for clinically significant bipolar disorder (one also with significant anxiety) and were excluded from the study. All remaining persons were offered enrollment and accepted, for a total of 23 persons enrolled for depression.

### Summary of Noncommunicable Disease Care Cascade

Out of 580 persons approached, the COMBINE enrollment process enrolled 23 persons with depression (4.0%) and 37 persons with hypertension (6.4%), none of whom met criteria for both conditions. However, the number of persons who initially screened positive for each condition was greater, with gradual attrition due to confirmatory testing; loss to follow-up; and screening errors. For instance 123 of 579 persons with a blood pressure check (21.2%) had an initial reading consistent with hypertension (including hypertensive urgency). However, only 97 were rechecked. And among those that were, only 58 were confirmed positive; 54 told they were positive; 45 linked to CHPS care (with 17 others), and only 33 had confirmed disease and were enrolled (with 4 others).

Similarly, though 159 of 574 persons with a home PHQ-2 scored 1 or more (27.7%), only 129 of them received a PHQ-9, and of these only 36 met study criteria, even though 84 (14.5% of the 580 approached) had at least mild depression. Further, of the 123 persons with an initial home blood pressure consistent with hypertension, 17 (10.1%) had a PHQ-9 consistent with depression, including 5 with moderate depression, and 10 (17.2%) of the 58 who met full study criteria for hypertension at home had depression, though only 1 had moderate depression. But among the 89 who linked to CHPS care - for hypertension and/or depression - only four met home criteria for both. Ultimately this person was excluded from the trial due to comorbid bipolar disorder. Therefore, no one enrolled met criteria for both conditions.

### Participant Demographics

The baseline demographic status of the 60 enrolled persons appears in Table 1. Persons with hypertension were significantly older on average (7.9 years) than those with depression (p < 0.01). Persons with hypertension reported exercising 194 more minutes per week, having an average of 1.1 more children per household, being 1.76 times more likely to report alcohol use and 2.51 times more likely to report tobacco use than persons with depression. However, only the difference in the number of children was statistically significant.

**Table 1:** Demographics of Enrolled Participants.

| Variable | Hypertension, N = 37<br>Mean (SD) | Depression, N = 23<br>Mean (SD) | p-value (for no difference) |
| --- | --- | --- | --- |
| Age in years (95% CI) | 54.2 (50.6-57.8) | 46.3 (42.5-50.2) | 0.00470* |
| Gender: N (%) |  |  |  |
| Male | 8 (21.6%) | 2 (8.7%) | 0.191** |
| Female | 29 (78.4%) | 21 (91.3%) |  |
| Disease Severity |  |  |  |
| Mean baseline BP or | 149/91 mm Hg | 13.3 (12.2-14.5) | N/A |
| PHQ-9 (95% CI) | (146-152/89-94) |  |  |
| Physical Exercise |  |  |  |
| Minutes per week (95% CI) | 985.0 (463.1-1506.9) | 790.7 (314.1-1267.2) | 0.867*** |
| Tobacco & Alcohol: N (%) |  |  |  |
| Tobacco | 4 (10.8%) | 1 (4.3%) | 0.379** |
| Alcohol | 17 (45.9%) | 6 (26.1%) | 0.104** |
| Level of education: N (%) |  |  |  |
| None | 24 (64.9%) | 16 (69.6%) | 0.467** |
| Primary | 8 (21.6%) | 2 (8.7%) |  |
| Secondary | 2 (5.4%) | 1 (4.3%) |  |
| Tertiary | 0 (0%) | 0 (0%) |  |
| Vocational/Technical | 0 (0%) | 0 (0%) |  |
| Other | 3 (8.1%) | 4 (17.4%) |  |
| Occupation: N (%) |  |  |  |
| Unemployed | 1 (2.7%) | 1 (4.3%) | 0.863** |
| Public Servant | 0 (0%) | 0 (0%) |  |
| Civil Servant | 1 (2.7%) | 0 (0%) |  |
| Farmer | 30 (81.1%) | 19 (82.6%) |  |
| Artisan/craftsman | 0 (0%) | 0 (0%) |  |
| Trader/business | 5 (13.5%) | 3 (13.0%) |  |
| Military | 0 (0%) | 0 (0%) |  |
| Professional | 0 (0%) | 0 (0%) |  |
| Religion: N (%) |  |  |  |
| Traditional religion | 14 (37.8%) | 3 (13.0%) | 0.154** |
| Islam | 4 (10.8%) | 4 (17.4%) |  |
| Christian | 18 (48.6%) | 16 (69.6%) |  |
| No religion | 1 (2.7%) | 0 (0%) |  |
| Other Specify | 0 (0%) | 0 (0%) |  |
| Ethnicity: N (%) |  |  |  |
| Kassena | 16 (43.2%) | 13 (56.5%) | 0.317** |
| Nankani | 21 (56.8%) | 10 (43.4%) |  |
| Builsa | 0 (0%) | 0 (0%) |  |
| Other Specify | 0 (0%) | 0 (0%) |  |
| Marital status: N (%) |  |  |  |
| Married/Living with a | 26 (70.3%) | 12 (52.2%) | 0.302** |
| partner |  |  |  |
| Divorce | 0 (0%) | 0 (0%) |  |
| Separated | 0 (0%) | 1 (4.3%) |  |
| Widowed | 10 (27%) | 8 (34.8%) |  |
| Never married | 1 (2.7%) | 2 (8.7%) |  |
| Type of marriage: N (%) |  |  |  |
| Monogamous | 20 (54.1%) | 7 (30.4%) | 0.148** |
| Polygamous | 13 (35.1%) | 9 (39.1%) |  |
| NA | 4 (10.8%) | 6 (27%) |  |
| Unknown | 0 | 1 |  |
| Number of Children<br>(95% CI) | 5.0 (4.4-5.6) | 3.9 (3.3-4.6) | 0.011* |

\*T-test
\*\*Chi-squared test
\*\*\*Wilcoxon rank-sum test

Participants in both groups were mostly female; middle-aged; had little to no formal education; and worked largely in farming. Persons from the Kassena and Nankani ethnic groups were almost equally represented. Participants were most commonly Christian, but with significant proportions adhering to traditional religions or Islam. Most were married, often in a polygamous marriage. The mean baseline blood pressure among persons enrolled for hypertension was 149/91, and the mean PHQ-9 score for those enrolled for depression was 13.3.

### Retention and Clinical Outcomes

Primary outcome data appear in Table 2. Among persons with hypertension, the mean blood pressure dropped by 26.7 mm Hg systolic and 16.0 mm Hg diastolic in the first 90 days. None of the 37 enrolled persons were lost to follow-up. At 180 days, all 37 persons again remained in follow-up, and systolic and diastolic blood pressures were not significantly different from the 90- day result. 5 and 3 persons respectively did not achieve blood pressure control (under 140 mm Hg systolic and under 90 mm Hg diastolic). At 270 days, one person left the study due to signs of stroke and was escalated to health centre care, and at 360 days, two more participants left due to acute medical complaints while one migrated away from the study zone. However, 34 persons achieved blood pressure control at 270 days, and at 360 days, 33 persons (100%) had achieved blood pressure control. In each case, systolic and diastolic blood pressure results were not significantly changed from day 90. However, relative to baseline blood pressure, all results were significant. There was no significant difference between genders for either condition: at baseline, men had a BP of 144/82 and women a BP of 151/91. In men with hypertension, BP decreased to 126/76 at 90 days and 120/77 at 1 year; for women the results were 121/75 and 121/75 at both time points. There was no significant difference in blood pressure between genders at either timepoint.

**Table 2:** Blood Pressure and Depression Treatment, Retention, and Control from 90-360 days^1^.

|  | Baseline | 90 days | 180 days | 270 days | 360 days |
| --- | --- | --- | --- | --- | --- |
| **Hypertension** | N = 37 | N = 37 | N = 37 | N = 36 | N = 33 |
| **Mean Systolic BP (95% CI)** | 149.1<br>(146.0-152.3) | 122.4<br>(117.5-127.4)<br>($p<.0001$)*** | 124.2<br>(120.5-127.9)<br>($p<.0001$)*** | 122.1 (118.6-125.6)<br>($p<.0001$)** | 120.6 (117.1-124.1)<br>($p<.0001$)*** |
| **Mean Diastolic BP (95% CI)** | 91.4 (89.2-93.6) | 75.4 (72.8-77.9)<br>($p<.0001$)** | 78.1 (75.8-80.3)<br>($p<.0001$)** | 76.2 (73.7-78.8)<br>($p<.0001$)** | 75.1 (71.9-78.2)<br>($p<.0001$)** |
| **Proportion Controlled (<140 mm Hg and < 90 mm Hg) and N** | 0% (N=35)<sup>2</sup> | 86% (N=32) | 92% (N=34) | 94% (N=34) | 100% (N=33) |
| **Proportion reporting tobacco use and N** | 10.8% (N=4) | 8.1% (N=3)<br>($p=0.001$)* | n/a | n/a | n/a |

<sup>1</sup> All $p$ -values for no difference between reported result and baseline.
<sup>2</sup> Two participants did not receive a baseline blood pressure check.

|  |  |  |  |  |  |
| --- | --- | --- | --- | --- | --- |
| **Proportion reporting alcohol use and N** | 45.9% (N=17) | 26.1% (N=6)<br>(p = 0.153)* | n/a | n/a | n/a |
| **Mean physical exercise minutes per week (95% CI)** | 985.0 (463.1-1506.9) | 896.5 (528.6-1264.5)<br>(p=0.537)*** | n/a | n/a | n/a |
| **Depression** | N = 23 | N = 20 | N = 19 | N = 18 | N = 18 |
| **Mean PHQ-9 (95% CI)** | 13.3 (12.2-14.5) | 2.0 (0.6-3.3)<br>(p<.0001)*** | 1.9 (0.9-3.0)<br>(p<.0001)** | 1.2 (0.4-2.0)<br>(p<.0001)*** | 1.4 (0.5-2.3)<br>(p<.0001)*** |
| **Proportion Controlled (PHQ-9 < 10)** | 0% (N=23) | 95%<br>(N = 19) | 100%<br>(N = 17)<sup>3</sup> | 100%<br>(N = 17)<sup>4</sup> | 100%<br>(N = 18) |
| **Proportion reporting tobacco use and N** | 4.3% (N=1) | 0% (N=0)<br>(p= n/a) | n/a | n/a | n/a |
| **Proportion reporting alcohol use and N** | 26.1% (N=6) | 10% (N=2)<br>(p=0.01)* | n/a | n/a | n/a |
| **Mean physical exercise minutes per week (95% CI)** | 790.7 (314.1-1267.2) | 696.6 (356.6-1036.5)<br>(p=0.9358) | n/a | n/a | n/a |
| **Total Proportion Retained and Controlled** | 0/60<br>(0% of 100%) | 51/57<br>(89% of 95%) | 51/54<br>(94% of 90%) | 51/53<br>(96% of 88%) | 51/51<br>(100% of 85%) |

\*T-test
\*\*Chi-squared test
\*\*\*Wilcoxon signed-rank test

Among persons with depression, the mean PHQ-9 score dropped by 11.3 points from baseline to 90 days, with 19 of 20 persons achieving depression control (PHQ-9 under 5). The rest of the patients migrated out of the study area. At 180 days, PHQ-9 score did not significantly change relative to 90 days - from 2.0 to 1.9. Two PHQ scores were not captured, and one participant was escalated from the study due to alcohol misuse requiring hospitalization. At 270 days, there one person migrated out of the study area. All persons assessed in the study at 180, 270, and 360 days (18 persons of 23 enrolled) achieved PHQ-9 control. None was given any antidepressant medication, as all persons remaining in the trial achieved the PHQ-9 endpoint after two visits. Mean baseline PHQ-9 scores were 12.5 and 13.4 for men and women respectively. Men with depression had a mean PHQ-9 of zero at 90 days and at 1 year (with 2 and 1 participants respectively) whereas women’s mean PHQ-9 was 2.17 at 90 days and 1.47 at 1 year. There was no significant difference in PHQ-9 between genders at either timepoint.

Tobacco, alcohol, and exercise data were captured only at baseline and at 90 days (primary study endpoint). All the 5 persons reporting tobacco use at baseline expressed interest in quitting; 3 were still using tobacco at 90 days. Due to inconsistent unit measurements (e.g., cigarettes versus rolled tobacco), quantitative changes in tobacco use could not be measured. Among the 23 persons reporting alcohol use at baseline, 21 expressed interest in cutting back or quitting. 7 were still consuming alcohol at 90 days, and 3 reported new alcohol use - as with tobacco, unit discrepancies precluded quantitative measurement of change. Both of these results were statistically significant overall, at p of <0.0001 and 0.01 respectively. However, the change in alcohol status was not significant among persons with hypertension. 58 persons reported their physical exercise level at baseline, and 55 at 90 days. Among both persons with hypertension and depression, the mean physical exercise level decreased (from 908 to 827 minutes weekly) but neither shift was statistically significant. ARMS data appears only for persons with hypertension, and only at 90 days. This result (not listed) was 15.46.

## Discussion

This pilot intervention demonstrated that Ghana’s CHPS primary healthcare initiative - which has achieved marked improvements in healthcare access and mortality for infectious, pediatric, and perinatal conditions - can treat two of the world’s leading noncommunicable diseases with strong efficacy across the care cascade of care linkage, retention, and control.^63,64^ 100% of eligible persons agreed to link to the intervention, compared to 82% in the prior CHPS-led hypertension TASSH trial.^25^ COMBINE retained 85% of enrolled persons in care at 1 year, the same result as TASSH, and significantly greater than the 25% one-year retention in the CHPS-led ComHIP hypertension trial (in which CHNs linked hypertensive persons to CVD nurses or physicians).^25,65^ COMBINE hypertension control rates were also comparable to these prior studies: 86% at 90 days and 100% at one year, relative to 55% for TASSH at 1 year and 72% for ComHIP.^25,65^ The mean absolute change in systolic blood pressure (28.5 mm Hg) was superior to both interventions (-20.4 mm Hg for TASSH and -12.2 mm Hg for ComHIP).^25,65^ With respect to depression, 1 year retention and PHQ-9 score change for persons were also comparable to similar interventions, such as the Friendship Bench program (87% retained at 90 days for COMBINE, 88% for Friendship Bench; mean PHQ-9 changes of 11.4 for COMBINE and 6.4 for Friendship Bench at 180 days.^66,67^ These results achieve the “80-80-80” goal proposed to define effective global control of hypertension,^68^ (now adopted by the Pan-American Health Organization),^69^ but also apply it to depression.

The COMBINE intervention also leverages CHPS’ radical task-shifting strategy more aggressively than prior efforts at hypertension control through Ghana’s public sector. TASSH and ComHIP leveraged CHPS nurses to detect hypertension proactively in rural villages, but linked suspected cases to nurses and physicians at health centres rather than treating them directly. These centres are part of the primary care infrastructure, but still inaccessible to many rural Ghanaians.^21,32,33^ Mental health interventions in Ghana’s public sector, such as the pilot District Mental Healthcare Plans,^70^ do not yet leverage CHPS directly, despite calls to use this resource for mental health screening and treatment.^35,71^ Providing CHPS-level prevention, testing, diagnosis and treatment for uncomplicated non-communicable diseases in Ghana could markedly improve all levels of the fragile care cascade.^11,39,64^

In addition to integrating with CHPS itself, COMBINE also integrates mental health care with NCD control, a priority combination for Ghana’s Ministry of Health^72^ but also for global health policy, as reflected in the 2025 Political Declaration on Noncommunicable Disease Control.^73^ Task-shifting is widely recognized as an effective yet underused strategy for both hypertension and depression control in LMICs, especially if integrated with treatment of other comorbid chronic diseases.^74-77^ but few NCD task-shifting proposals in Ghana or elsewhere propose to integrate the two at the community level.^28,29,78,79,80^ The precise approach through which to integrate decentralized care for these leading causes of death and disability remains poorly understood, warranting further evaluation at scale within robust established national models for community health care such as CHPS.^23,81^

### Strengths and Limitations

Although leveraging an existing village-level cadre of primary care practitioners and health counselors for integrated NCD and mental health care may improve COMBINE’s feasibility and scalability, the pilot structure of the intervention to date limits its applicability to the Ghanaian and global policy interventions above. The trial was not designed with a control group, nor with sufficient power to detect a clinically significant change in blood pressure or depression severity relative to the control group or to baseline. The under-representation of male gender further limited analysis, especially in men with depression (n = 2). The trial did not fully interrogate behavioral mediators of the intervention, such as medication adherence (measured once), program satisfaction, or quality of life.^82,83^ Moreover, it did not evaluate the cost-effectiveness of the intervention, nor its perceived scalability within the Ghana Health Service. Lastly, because no participant screened positive for both hypertension and depression, the intervention does not provide any data on multimorbidity per se. However, as noted above, multiple individuals screened positive for both conditions (e.g., 10 of 58 persons with an average home blood pressure in the study range had a PHQ-9 of 5 or more), such that steps to simplify the screening process or lower the depression or hypertension diagnostic cutoff could aid identification and recruitment of persons with multiple comorbid NCDs in a subsequent randomized trial.

## Conclusions

The COMBINE pilot intervention demonstrates substantial promise in diagnosing and treating two of the leading global causes of noncommunicable disease, by leveraging front-line nonphysician health workers from one of the world’s leading rural healthcare initiatives. Scale-up of this intervention to a powered, cluster-randomized, controlled trial could evaluate both the efficacy and implementation feasibility of COMBINE, as well as its cost-effectiveness and mechanisms of action. If successful in targeting the underlying behavioral determinants of multiple chronic conditions, the COMBINE care model could expand not only across Ghana and similar health systems, but also across other NCDs such as diabetes or anxiety. This approach could provide a novel pathway for integrated, proactive NCD testing and treatment in underserved rural communities worldwide.

## Data Availability

All data produced in the present study are available upon reasonable request to the authors.

## Acknowledgements

We thank the staff, research team, and data collection team of the Navrongo Health Research Centre; our colleagues at the Ghana Health Service; and the patients and participants who engaged in this study, as well as the Arnhold Institute for Global Health for their support.

## Footnotes

1 All p-values for no difference between reported result and baseline.

2 Two participants did not receive a baseline blood pressure check.

3 Two participants did not receive a 180-day PHQ-9 check.

4 One participant did not receive a 270-day PHQ-9 check.

